# Risk of secondary infection waves of COVID-19 in an insular region: the case of the Balearic Islands, Spain

**DOI:** 10.1101/2020.05.03.20089623

**Authors:** Víctor M. Eguíluz, Juan Fernández-Gracia, Jorge P. Rodríguez, Juan M. Pericás, Carlos Melián

**Affiliations:** Instituto de Física Interdisciplinar y Sistemas Complejos IFISC (CSIC-UIB), E07122 Palma de Mallorca, Spain; ISI Foundation, Turin, Italy; Infectious Disease Department, Hospital Clínic de Barcelona, Barcelona, Spain.; Vall d’Hebron Institute for Research (VHIR), Barcelona, Spain; Department of Fish Ecology and Evolution, EAWAG Swiss Federal Institute of Aquatic Science and Technology, Centre of Ecology, Evolution and Biogeochemistry, Seestrasse 79, CH-6047, Kastanienbaum, Switzerland; Institute of Ecology and Evolution, Aquatic Ecology, University of Bern, Baltzerstrasse 6, CH-3012, Bern, Switzerland

**Keywords:** COVID-19, epidemic projection, secondary outbreaks, computational modeling

## Abstract

The Spanish government declared the lockdown on March 14th, 2020 to tackle the fast-spreading of COVID-19. As a consequence the Balearic Islands remained almost fully isolated due to the closing of airports and ports, These isolation measures and the home-based confinement have led to a low prevalence of COVID-19 in this region. We propose a compartmental model for the spread of COVID-19 including five compartments (Susceptible, Latent, Infected, Diseased, and Recovered), and the mobility between municipalities. The model parameters are calibrated with the temporal series of confirmed cases provided by the Spanish Ministry of Health. After calibration, the proposed model captures the trend of the official confirmed cases before and after the lockdown. We show that the estimated number of cases depends strongly on the initial dates of the local outbreak onset and the number of imported cases before the lockdown. Our estimations indicate that the population has not reached the level of herd immunization necessary to prevent future outbreaks. While the low prevalence, in comparison to mainland Spain, has prevented the saturation of the health system, this low prevalence translates into low immunization rates, therefore facilitating the propagation of new outbreaks that could lead to secondary waves of COVID-19 in the region. These findings warn about scenarios regarding after-lockdown-policies and the risk of second outbreaks, emphasize the need for widespread testing, and could potentially be extrapolated to other insular and continental regions.

## Introduction

The rapid propagation of the new COVID-19 pandemic requires timely responses, including the alignment of evidence generation by scientists and decision-making by policy stakeholders. As of the current date, several mathematical models have been developed to help policy-making in a wide arrange of interventions in various countries, encompassing from testing strategies to lockdown measures (*1*–*6*). Although modeling pandemics is not without flaws, and its predicted scenarios cannot be uncritically adopted and therefore directly translate into policy (7), modeling can be a valuable support tool to guide policy when assessed in an integrated way.

Recent studies have dealt with the possibility of a second-wave of COVID-19 after the retirement of lockdown and confinement measures in China (1–2). Recently, the value of restrictive social distancing measures has been recommended in Italy (8). The analysis of data from closed confinements such as sea cruises allows us to address some key questions regarding the risk of second waves in an environment without external perturbations (9,10). The study of the evolution of the pathogen in islands offers an opportunity to learn how the propagation occurs, and how the mobility restrictions are shaping the propagation in relatively isolated areas, either due to transport lockdowns implemented to contain COVID-19 dissemination or because of their geographical conditions.

The Balearic Islands archipelago is composed of four inhabited islands in the Mediterranean Sea, i.e Majorca, Menorca, Ibiza, and Formentera, with a total population of 1,095,426, as per 2011 data (11). The main economic activity is tourism with principal connections to the UK and Germany. The first reported case in Spain was identified in the Canary Islands on January 31st, while in the Balearic Islands the first case (second in Spain) was confirmed on February 9th. He was a British citizen resident in Majorca who had been in contact with an infected person with SARS-CoV-2 during a stay in France from January 25-29. In Spain, the schools were closed on March 16th and the lockdown was implemented at the national scale from March 17th. As of April 11th, the number of infected in the Balearic Islands was 140 per 100,000 inhabitants to be compared with 354 in Spain (data updated with values of April 11th, 2020) (12). The lockdown of the Balearic Islands includes the closing of airports and ports for passengers, rendering the archipelago a virtually closed system. In this regard, archipelagos are “living laboratories” suggesting insights about the ecology and evolution of infectious diseases and offering unique experimental testing protocols to reduce or eliminate the diseases not only in the islands but potentially across the world (13). Thus, the Balearic Islands present an opportunity to be used as a benchmark to explore how isolation and after-lockdown measures impact secondary COVID-19 waves.

COVID-19 has a particular structure in the timings of the disease that make it particularly dangerous in terms of a silent spreading potential. First, the incubation period, i.e the time since infection to symptom onset, is relatively large around 5.2 days (95% confidence interval [CI], 4.1 to 7.0) (14). This itself is a driver of the predictability of the spatiotemporal patterns to expect from this disease (15). Furthermore, the latent period, i.e the time since infection to the start of being infectious, does not align completely with the incubation period (4). This leaves a period of presymptomatic infectivity, that increases R_0_ through silent spreading, as not even the carrier might be aware of its own infectivity (16). The relative effectiveness of different non-pharmaceutical interventions will depend critically on the relation of those times (incubation and latent period) (17). Other related periods that shape the dynamics of the outbreaks are the generation interval (time between infection of infector-infectee pairs) (18) and the serial interval (the time between symptom onsets of an infector-infectee pair), which has also been used to estimate viral shedding dynamics for COVID-19 (4).

We aimed to study the dissemination of COVID-19 in a quasi-isolated system through a compartmental model that included, besides the susceptible (S), diseased (D) and recovered (R) compartments, an exposed (E) compartment, and a pre-symptomatic (I) infective compartment to account for the incubation period, as the times of transit between the latter two compartments are crucial for the modeling of COVID-19 (3, 4). Due to population size, we can implement an individual-based model where we consider each inhabitant as an individual in the model. In particular, first, we compare the results of an individual-based model tailored for the Balearic Islands and identify the parameter values that best fit the data. Second, we explore the likelihood of a second-wave scenario as a function of the initial date of the first imported case and the number of imported cases before the lockdown.

## Results

### Number of active infected cases

The best fit of the model to the confirmed cases, allows us to extract the transmission probabilities and also the scaling factor that captures the ratio between the estimated and the confirmed cases. For the scenario where the initial date was on Feb 7^th^, and a latent period of 2 days (*T*_lat_=2), an infective period of 4 days (*T*_inf_=4), and disease period of 12 days (*T*_dis_=12), the values of the infection probability leading to the best fit are β_1_=0.24, β_2_=0.12, β_3_=0.016, and β_4_=0.036 (Figure 1). This translates into an initial basic reproductive number *R*_0_=3.84. The fitting of the data also informs us that the correction factor is 0.054, that is, that the confirmed cases are 5.4% of the model estimates. At the same time, we obtain that the percentage of added healed cases and fatalities according to the official sources the model estimate are 5.3% of the confirmed cases. For the other values of the latent, infective, and disease periods, we obtain similar accuracy, given by χ^2^ and similar scaling factors (Table S1). The scaling factor, which gives the fraction of the model estimates, that corresponds to the confirmed case, increases to 10% in the case of *T*_lat_=5, *T*_inf_=1, which is the case with less infected individuals in the model.

**Fig. 1.**
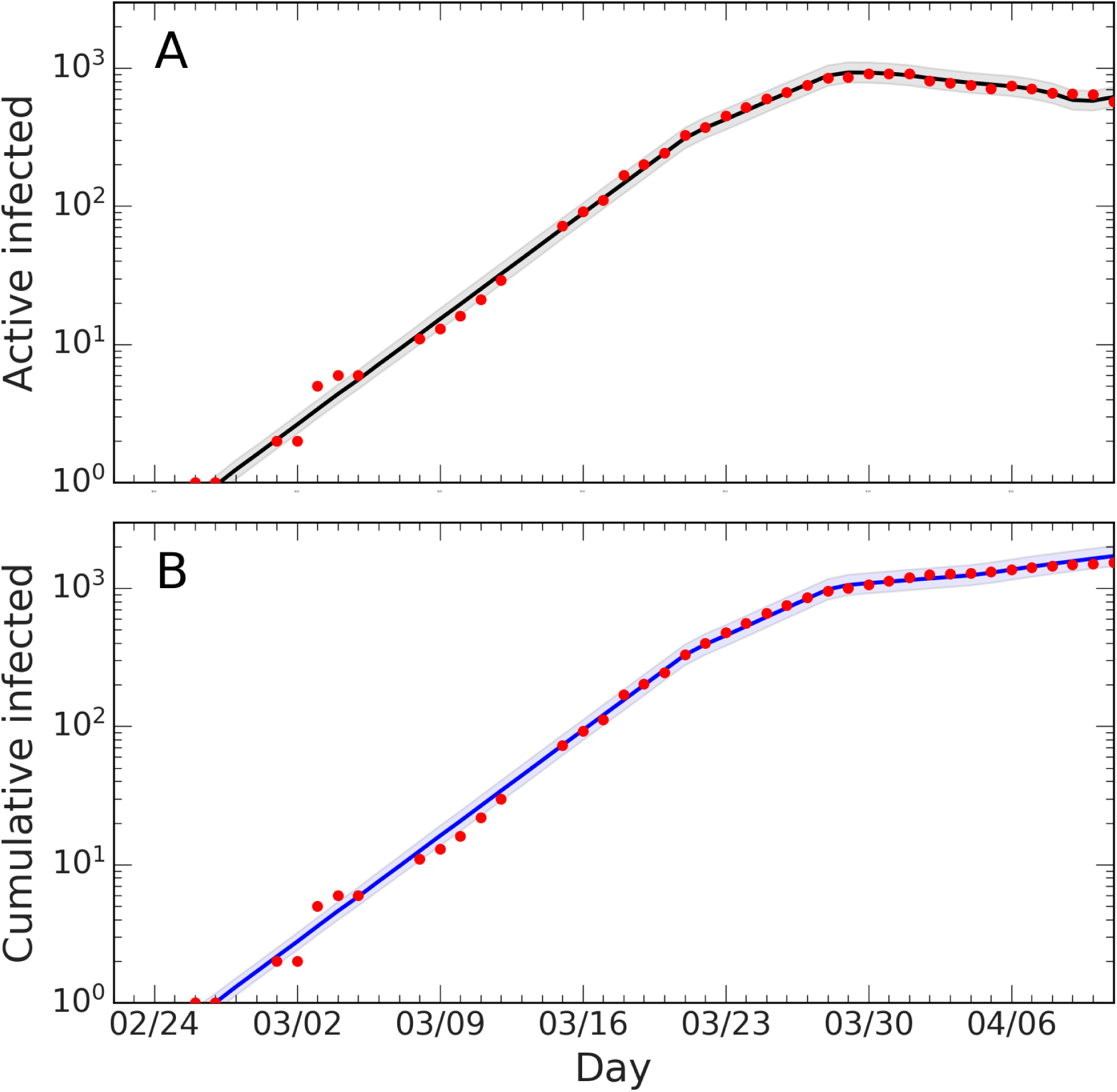
Active and cumulative infected time series in the Balearic Islands. Time evolution of the average number of (A) active infected cases (black line) and (B) accumulated number of cases with the best-fitted parameters, and confidence interval CI 90% (grey area). Red dots represent the data provided by the Ministry of Health of Spain, and the solid lines depict the model with the fitted parameters *T*_lat_ = 2, *T*_inf_ = 4, *T*_dis_ = 12, *β*_1_ = 0.24, *β*_2_ = 0.12, *β*_3_ = 0.016, *β*_4_ = 0.036, *β*_5_ = 0.

The introduction of a single imported infected case after the first wave has expired strongly correlates with the risk of a secondary wave (Figure 2). The intensity and duration of the second wave depend on specific values capturing the conditions applicable when newly infected cases appear, e.g the transmission probability, which depends on the habits of the population, hygiene, and social distancing, and mobility restriction. Qualitatively similar results were obtained for the other set of values. The peak of the second wave is very sensitive to the date of the first exposure. If it happened on January 28^th^, the intensity of the second peak is less pronounced and similar to the one for the first peak, in contrast to the case of a more recent exposure, when the second peak can be more than one decade larger the first peak.

**Fig. 2.**
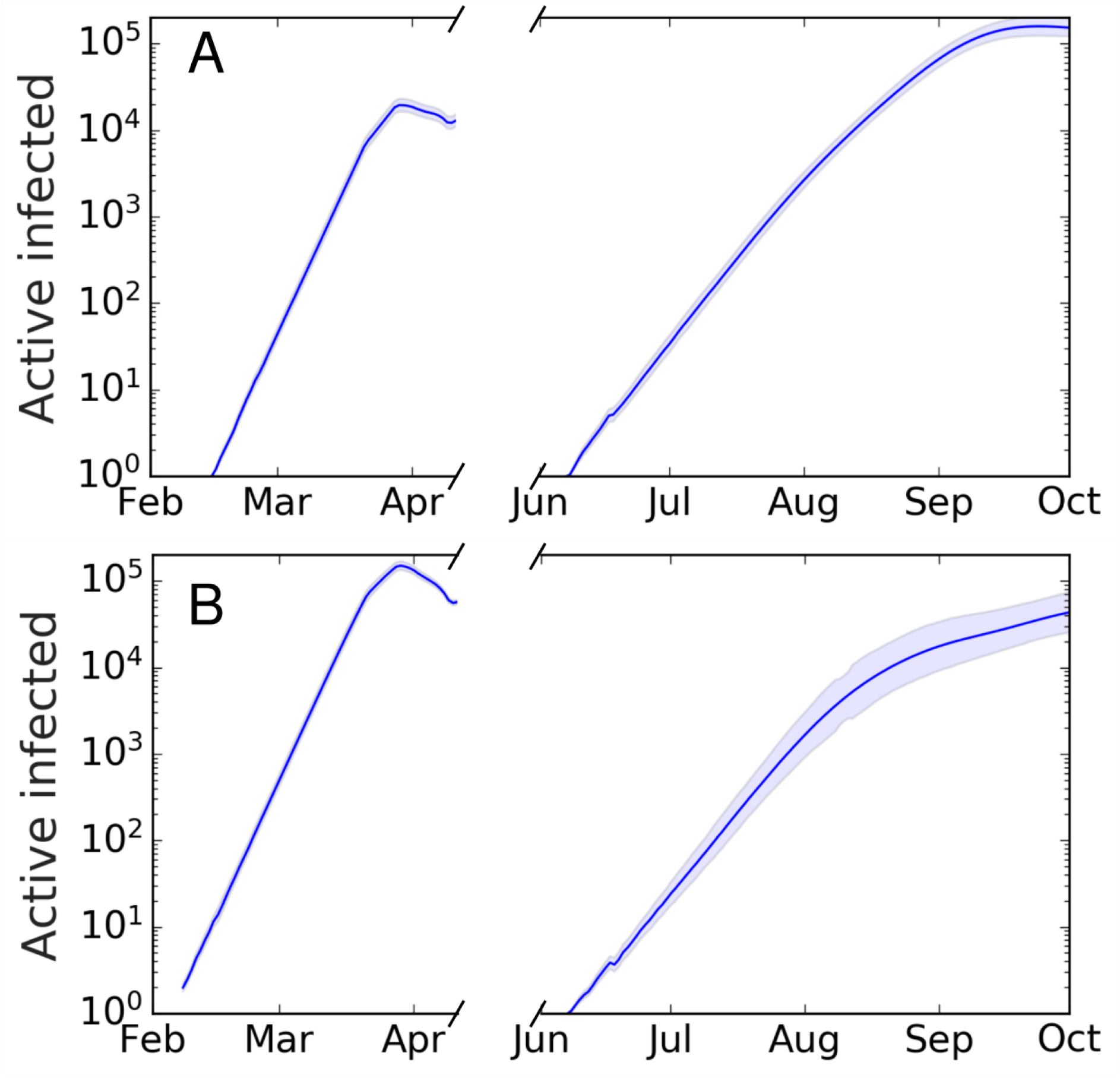
Secondary outbreak appearing after the home-based confinement is removed. The lockdown is removed and the parameter values are as during the week of March 16^th^-22^nd^. (A) First infected case is on February 7^th^, the number of infected in the first wave has not reached herd immunization and a second wave is triggered by a single infected case. The intensity of the second wave is one order of magnitude larger than the first. (B) First infected case is on January 28^th^, the number of infected in the first wave has reached a larger fraction of the population and the intensity of the second wave is, in the scenario, smaller than the first. Average over 100 realizations.

### Herd immunization estimates

Assuming recovered individuals get immunity, to estimate whether the Balearic Islands have reached herd immunization, we explored the estimated number of infected under two immunization scenarios based on the date of the first infection (Figure 3A) and the number of imported cases before air and maritime transport lockdown (Figure 3B). Firstly, we analyzed how the estimation of infected individuals is sensitive to the date of the first infection. We explored the time range of the first infection from January 28^th^ (which corresponds to the stay in France before returning to Majorca) to Feb 7^th^ (which corresponds to 30 days before the 10th confirmed case). Secondly, we explored the estimates under the assumption that more than one imported case could have gone unnoticed into the Balearic Islands before the closing of the airports. Depending on these two parameters, the range of immunization spans from 3% (for one initial infected on February 7th) to 64% (for 20 initial infected cases on January 28th). With the assumption of immunity after recovery, the achievement of herd immunization in the population is very sensitive to the date of the first infection and the number of imported cases before air and maritime transport lockdown. The interpretation of herd immunization indicates that if infected individuals become immune, then 20% of herd immunization prevents the spreading of reproductive numbers smaller than 1.25. Assuming that the first case was exposed to the infection during his stay in France in the last days of January, the percentage of the population that was infected can be as high as 50%, which could prevent a high second peak, for values of the basic reproductive number below 2. Conversely, if the first case was infected 30 days before 11 confirmed cases were reported in the Balearic Islands, the percentage of infected individuals could lower to less than 10%, therefore falling far from potential herd immunity (only for values of the basic reproductive number below 1.1). The relation between the number of initial infected cases, the date of the first infection, the number of cases, and the number of confirmed cases is further explored in the Supplementary Material.

**Fig. 3.**
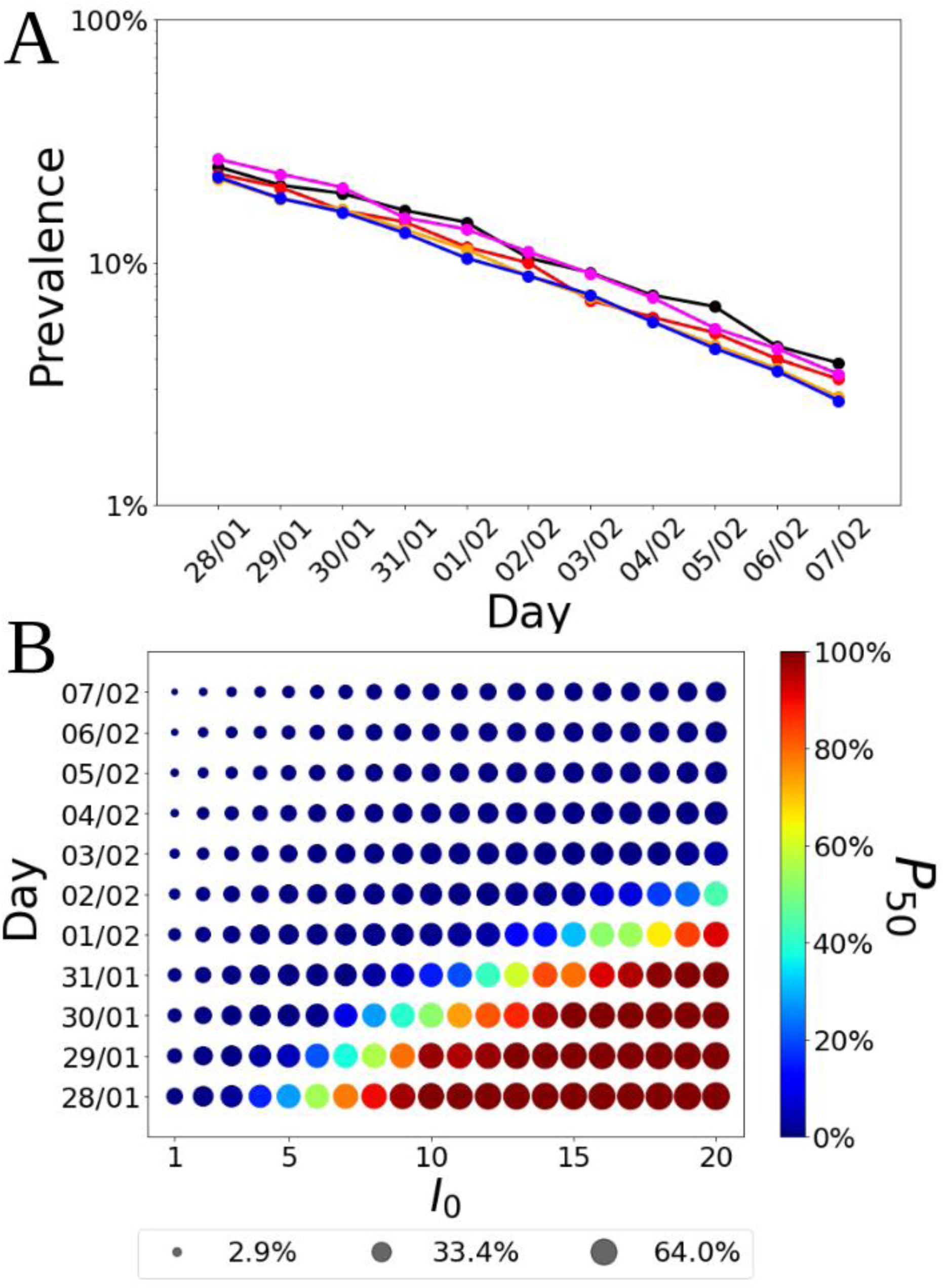
Fraction of infected individuals (in logarithmic scale) as a function of the date of the first infection. (A) Each line corresponds to the following parameter values(*T*_lat_, *T*_inf_)=(1,5) (black), (2,4) (red), (3,33 (orange), (4,2) (magenta), and (5,1) (blue). (B) Fraction of infected cases as a function of the first infection and the initial number of infected individuals. The radius of the symbol is proportional to the fraction of infected cases while the color indicates the probability that a realization of the model reaches at least 50% of infected cases in the population, *P_50_*.

## Discussion

Our study shows that a model including five compartments together with information on mobility between municipalities can be used to capture the spread of the epidemics in a closed community. The validation of the model with the official data allowed us to obtain the parameters that best fitted the data. Once the model was validated, we extracted an estimation of the number of the total infected in the Balearic Islands that indicates, assuming immunization after recovery, that these figures would reach the herd immunization threshold depending critically on the date of the first infection and the initial number of seeds, being herd immunization achievement more likely for an initial date before January 31st and number of initial infected above 10. Our exploration of the forecasted scenario of a newly infected individual entering the community after the lockdown confirmed that the number of potential cases widely varies according to the initial date of infection, which correlates with the percentages of immunity. Although we cannot determine with precision the start of the infection in the Balearic Islands, the model suggests that the Balearic Island population is below the herd immunization threshold and thus, also susceptible to new outbreaks depending on how immunity is acquired and how the mobility restrictions are further implemented. In particular mobility and transmission probability, which depends on the general use of masks and hygiene protocols by the population, might alter the attack rate.

Focusing on second waves in insulated areas during the COVID-19 pandemic is of great value to analyze the spreading and containment of infectious diseases, where the lockdown of islands constitutes a paradigmatic scenario, with the potential to be applied to continental regions (13). The use of modeling tools is a complement to field studies that can be used to anticipate the progress of a pandemic and thus help health authorities to make decisions. In the case of the Balearic Islands, there are two foremost advantages in terms of model precision. First, since the incidence during the first peak was relatively low and hospital capacity including ICU beds was not overpassed, the forecasted scenario of a second wave presenting with more intensity is more feasible than in other areas. Second, the relatively small size of the Balearic Islands and the organization of health and epidemiological surveillance systems make the official accounts of reported cases more reliable than in other areas were due to low rates of testing, overloaded hospitals, and lack of centralized data collection hampered the initial estimates.

The implications of the forecasted potential second wave yielded by our model for an insular territory can be useful also for other areas that either naturally geographically isolated or closed to external perturbations due to strict lockdowns. According to our results, the date of the first infection and the import of cases while the airports and ports were open appear to be key to assess the likelihood and intensity of future waves and outbreaks. Knowing the approximate date of infection of the first reported case in an outbreak proved critical to estimate the current and foreseen number of cases. Whether a second wave occurs and the intensity of the peak strongly depended on the date of the first infection, as the number of infected cases grows exponentially, but also on the number of imported cases, which contribute additively to the number of cases, and also on the real herd immunization. Our estimates rely on calculations assuming conditions far from the behavior of the population, and on the habits, for example, regarding hygiene, the use of masks, and social distancing, of the population after the lockdown is relaxed.

Our model is an individual-based model for which, due to the population size, we identify each inhabitant with an individual in the simulation model. This approach is different from other models considering pan-mixing and ordinary differential equations (8). Other approaches implement recurrent mobility (3, 19, 20), which selects the individuals that perform the commuting randomly at each iteration step, thus increasing the mixing in the complete population. Our approach assigns a residence and a working municipality as initial condition and these locations remain unchanged during the time evolution for the model. Our implementation assumes that the same person commutes between two locations and thus it has to be fixed initially in the model. A random selection at each day will increase the number of effective connections, which could be compensated by a reduction in the transmission probability. We believe this approach is more comprehensive and better captures the reality of commuting under home confinement conditions, which essentially limit mobility from households to workplaces for those individuals that cannot work at distance or are not exempted from any work under the regulations of each country, while the rest of the population are not supposed to move from the vicinity of their households and even so just for justified reasons as basic food supply. We use here a stochastic approach similar to other works (19, 21) which lets us compute confidence intervals even for single combinations of the parameters instead of deterministic ordinary differential equations (8, 22, 23) or discrete-time dynamic equations (3). We also use a fixed time for the transit through the E, I, and D compartments, *T*_lat_, *T*_inf_, and *T*_D_ respectively. We believe this approach is more realistic than an approach based on rates, where individuals transit the compartments at a given rate, giving rise to exponentially distributed times of transit through the compartments. In this case, infected will have the opportunity to be infectious immediately, or to transit the I compartment also immediately, bringing the start of secondary infections closer to the time they were infected for many individuals. A similar effect happens with the length of the disease (time in the D compartment), having then individuals that immediately recover.

The model also has several limitations. First, as it is constructed for fitting the global numbers of infected patients, it is missing finer structure, needed for the evaluation of risks of subpopulations that are differently exposed to the virus or have different outcomes, such as the population of elderly people or health workers. Second, for COVID-19 there is evidence of three main transmission channels, namely direct contact with an infected individual with symptoms (14), contacts with an asymptomatic individual (24, 25), and environmental transmission (26). The present model takes into account the first two modes of transmission, but not the environmental one explicitly, although probably the fitting has assigned part of this transmission to the processes included in our model. Therefore, there is not a direct way of measuring the effect of interventions to reduce environmental transmission. Third, the model also considers asymptomatic and symptomatic individuals to be infectious in the same way, although the viral shedding in asymptomatic individuals is indeed lower (5). This can have a mild impact on the number of infected individuals that the model predicts. Fourth, the model assumes that the mobility restrictions are applied in the same way to all of the agents in the system and thus is lacking the fact that symptomatic infected individuals will modify their mobility drastically, either if they are quarantined at home or admitted to a hospital. We are therefore overestimating mobility, but this is probably passed to the infectivity in the fitting procedure. Fifth, the model also takes fixed times to transit through the E, I, and D compartments (*T*_lat_, *T*_inf_ and *T*_D_, respectively), which is artificial. More refined models would take this transit times from specified distributions matching the parameters of the disease (21, 23). While this will render the model a more realistic approach, we believe that fixing the times is a good compromise between using rates for transiting the compartments and implementing distributions for those times, as it already captures the delays induced by these particular timings of the infection. Finally, the model also assumes that individuals are granted immunity to the virus, at least for the timescales explored here.

In conclusion, the risk of secondary infection waves should be comprehensively and cautiously addressed before removing confinement measures. Our study provides several relevant findings that could be useful to support policy design at avoiding second waves once measures to return to the societal usual activities start to be applied. First, the isolation of asymptomatic individuals that tested positive for COVID-19 and close contacts to infected individuals during the prior two weeks might reduce the number of new infections after the establishment of the usual activity by preventing dissemination from asymptomatic carriers during the incubation period. This requires proper testing strategies tailored according to the estimated prevalence of infection, population density, the openness of the community, and other relevant factors. Second, contact tracing measures are crucial, and digital tools might enhance the identification of high-risk individuals to be tested or preemptively isolated (27). Yet, data privacy and other relevant ethical considerations should be carefully balanced when designing contact tracing in the community. Third, progressive return to the normal activity instead of an abrupt change will facilitate the monitoring of new cases and may avoid a sharp growth of the number of infected individuals, which is expected when herd immunity has not been reached. Further modeling studies on second-waves of COVID-19 are warranted to strengthen the knowledge on the best theoretical assumptions and data to be used to increase forecasting precision. In addition, these models should be validated through real-world data as these are collected during and after the pandemic.

## Material and Methods

### Data

Population data for the 67 municipalities in the Balearic Islands were taken from the Instituto Nacional de Estadística (INE, Spain), which gathers all the census data (11). The census also provides the commuting flows for people that, according to the registry, are living in one municipality and work in another. This allows assigning a living location and working location to each individual. For small municipalities, these commuting fluxes are not included. We avoid the isolation of these municipalities considering commuting flows of 10 people towards each of the neighboring municipalities and Palma, the capital of Balearic Islands.

Data for the active infected and accumulated infected cases are obtained from the Ministry of Health (12). In particular, the official reports provide data on the accumulated number of infected, healed, and deaths. The number of active infected cases is taken as the number of accumulated infected and subtracting deaths and healed. Unfortunately, the values for healed cases are only reported from March 22nd. Thus for the fitting, we only considered values starting from March 22nd.

### Model

The relatively small population size of the Balearic Islands allows us to develop an individual-based model. Each individual is placed in one of 67 municipalities according to the census (Figure S1). The mobility between municipalities is considered with commuting data from the 2011 census provided by the INE (11). For each simulation day, we consider two steps, one where each individual is located in its residence municipality, and a second step where each individual is placed in the working place. At each step, individuals can interact with any of the individuals placed at the same location. The locations assignment is made by randomly selecting a residence and a working place respecting the populations from the data. This assignment is done initially and such positions remain unchanged during the time evolution of the model.

The states of the individuals correspond to a SEIDR model: S, susceptible; E, exposed, corresponding to the latent period; I, infectious, corresponding to the presymptomatic infective period; D, diseased, corresponding to be infective with or without symptoms; and R, recovered. The transitions between these states are as follows, S becomes E in contact with an infected (I or DI) with probability β. After *T_lat_* (latent period) days, E becomes I; after *T_inf_* (presymptomatic infective period) days I becomes D, and after *T_dis_* (disease), D becomes R.

The values of *T*_lat_, *T*_inf_, and *T*_dis_ were obtained from the time evolution of the number of active infected and accumulated infected cases in the Balearic Islands. The lockdown was imposed in Spain on March 16th and the effect of the mobility restrictions can be identified on March 22nd. The 6 days in this period are reflected in the condition *T*_lat_ + *T*_inf_ (Figure S2A). Finally, from the data on the accumulated number of infected, the change in slope is observed on April 2nd, that is, *T*_dis_ = 12 days (Figure S2B).

To implement the mobility restrictions, we observe from the data that the accumulated number of infected shows a bending every 7 days approximately, which is in accordance with the beginning of the lockdown, and the restriction imposed on March 15th, and later corrected on March 22nd and March 29th. Thus the model has the freedom to adjust the infection probability every week after March 15th.

For a single day, the modeling proceeds as follows,

- 1. First, it considers the population in their residence location, for each municipality pairs of individuals in the same municipality are selected, say *i* and *j*. Then, *i* updates his/her state according to the dynamic rules. For each municipality *p, N_p_* pairs are chosen randomly where *N_p_* is the population size of the municipality *p*.
- 2. Second, we consider the individuals distributed in the municipalities of work. For each municipality *p*’, we chose *N_p_* pairs of random individuals working in the same municipality *p*’.
- 3. Resume from 1.

Thus, on average, in a day, each individual is updated once.

For calibration, the model is run exploring all the parameters:

- *T*_lat_ + *T*_inf_ = 6
- *T*_dis_ = 12

and β is explored in the range [0,1] in the following periods:

- β_1_ from the origin of the infection on February 9th to March 15^th^,
- β_2_ from March 16th to March 22nd,
- β_3_ from March 23th to March 29th,
- β_4_ from March 30th to April 5th,
- β_5_ from April 7th to April 11th

The total number of infected depends on the date of the first infection. Models assume that the beginning of the outbreak is typically 30 days before the day when 10 infections are recorded (28). In the case of the Balearic Islands, on March 8th, 11 confirmed cases were reported. The first case reported in the Balearic Islands corresponds to an imported infection notified on February 9th. Consequently, the beginning of the outbreak was set on February 7th, two days before the first infected was identified. Thus, we explore the date of the beginning of the infection between Jan 28th and Feb 7th.

### Model validation

The results of the model are validated with the official number of active infected and the accumulated number of infected cases between March 15^th^ and April 11th. As the official values do not take into account the non-tested asymptomatic and the diseased not consulting to the healthcare systems, we assume that the reported values are a proportion of the values obtained from the model. Then, to validate the model parameters we minimize χ^2^, χ^2^ = Σ (*α Y_i_* - *y_i_*)^2^, where α is a scale factor, that is, the ration between estimated and confirmed cases, *Y_i_* is the value obtained from the model in day *i*, and *y_i_* is the official value in day *i*. Due to the initial exponential growth of the epidemics, we calculate χ^2^ for the logarithm of the cases: χ^2^= Σ (log(*α Y_i_*) - log(*y_i_*))^2^. The minimization of χ^2^ leads to an optimal scale factor log(α^*^) =1/*n* Σ (log(*y_i_* / *Y_i_*)). For this value of α^*^, we finally calculate the optimal values. Our assumption implies that the scale factor should be similar to both the active and accumulated infected.

For each set of parameters, we report the χ^2^ of the model values of the number of active infected cases with respect to the official values, the correction fraction α_active_, and the 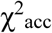 of the model values of the number of accumulated infected individuals with respect to the official vales and the correction fraction α_acc_. For each set of parameters, the best fit is considered as the one leading to the minimum χ^2^ Once the fitting values are determined, we calculate 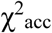 and α_acc_ For each set of periods (*T*_lat_, *T*_inf_, *T*_dis_), we explore the infection probabilities that minimize χ^2^ of the number of active infected cases. The value of χ^2^ and scale factors α^*^ of the best fits are shown in Table S1 and the estimated incidence in Table S2.

### Herd immunity assumptions

An approximation to the herd immunity threshold is given by 1-1/R_0_ (29), which for COVID-19 is expected to be between 29 and 74%, taking R_0_ between 1.4 and 3.9 (14,30). To explore whether the number of accumulated infections reach the herd immunity threshold and therefore avoidance of potential second waves is to be expected, we run the model for the same parameters leading to the best fit (29). After the system has relaxed to zero infection, we select a random susceptible from the populations and infected her. As we are interested in whether the epidemics will spread again, we use the transmission rate obtained at the beginning of the epidemics in the Balearic Islands, that is, before any restriction on mobility had been applied. We can expect that once the mobility restrictions have been removed, the transmission will be reduced in comparison to the initial values, especially due to an improvement in the hygiene of the population. This will affect how fast the COVID-19 will spread and the intensity of the wave. If the estimated number of infected is lower than the threshold for herd immunization, we assume that the COVID-19 will spread. In the Supplementary Material, we show how the data can be collapsed using a proper combination of the initial number of infected cases and the time of the first infection.

## Data Availability

Data is available as indicated in the main text.

## Acknowledgments

V.M.E. and J.F.G. acknowledge funding from the Ministry of Science and Innovation (Spain) and FEDER through project SPASIMM [FIS2016-80067-P (AEI/FEDER, UE)]. JFG acknowledges funding through the postdoc program of the University of the Balearic Islands.

## Authorship contributions

V.M.E. and J.F.G. designed the work, V.M.E. performed the analysis, V.M.E J.F.G J.P.R J.M.P and C.M. prepared the figures, tables, wrote the first draft and provided final approval to the manuscript. The corresponding author attests that all listed authors meet authorship criteria and that no others meeting the criteria have been omitted.

## References

1. K. Leung, J.T. Wu, D. Liu, G.M. Leung. First-wave COVID-19 transmissibility and severity in China outside Hubei after control measures, and second-wave scenario planning: a modeling impact assessment. Lancet 395, P1382–1393 (2020). DOI: https://doi.org/10.1016/S0140-6736(20)30746-7

2. Lei Zhang, Mingwang Shen, Xiaomeng Ma, Shu Su, Wenfeng Gong, Jing Wang, Yusha Tao, Zhuoru Zou, Rui Zhao, Joseph Lau, Wei Li, Feng Liu, Kai Ye, Youfa Wang, Guihua Zhuang, Christopher K Fairley. What is required to prevent a second major outbreak of SARS-CoV-2 upon lifting the quarantine of Wuhan city, China. *medRxiv* 2020.03.24.20042374 (2020); doi: 10.1101/2020.03.24.20042374. Available at https://www.medrxiv.org/content/10.1101/2020.03.24.20042374v1

3. Alex Arenas, Wesley Cota, Jesus Gomez-Gardeñes, Sergio Gómez, Clara Granell, Joan T. Matamalas, David Soriano-Panos, Benjamin Steinegger. A mathematical model for the spatiotemporal epidemic spreading of COVID19. *medRxiv* (2020). 03.21.20040022; doi: 10.1101/2020.03.21.20040022. Available at https://www.medrxiv.org/content/10.1101/2020.03.21.20040022v1

4. Xi He, Eric H. Y. Lau, Peng Wu, Xilong Deng, Jian Wang, Xinxin Hao, Yiu Chung Lau, Jessica Y. Wong, Yujuan Guan, Xinghua Tan, Xiaoneng Mo, Yanqing Chen, Baolin Liao, Weilie Chen, Fengyu Hu, Qing Zhang, Mingqiu Zhong, Yanrong Wu, Lingzhai Zhao, Fuchun Zhang, Benjamin J. Cowling, Fang Li and Gabriel M. Leung. Temporal dynamics in viral shedding and transmissibility of COVID-19. Nat Med (2020); doi: 10.1038/s41591-020-0869-5. Available at: https://rdcu.be/b3ADi

5. Neil M Ferguson, Daniel Laydon, Gemma Nedjati-Gilani, Natsuko Imai, Kylie Ainslie, Marc Baguelin, Sangeeta Bhatia, Adhiratha Boonyasiri, Zulma Cucunubá, Gina Cuomo-Dannenburg, Amy Dighe, Ilaria Dorigatti, Han Fu, Katy Gaythorpe, Will Green, Arran Hamlet, Wes Hinsley, Lucy C Okell, Sabine van Elsland, Hayley Thompson, Robert Verity, Erik Volz, Haowei Wang, Yuanrong Wang, Patrick GT Walker, Caroline Walters, Peter Winskill, Charles Whittaker, Christl A Donnelly, Steven Riley, Azra C Ghani.. Impact of non-pharmaceutical interventions (NPIs) to reduce COVID19 mortality and healthcare demand. Imperial College COVID-19 Response Team Report. (2020), March 16; doi: 10.25561/77482. Available at: https://www.imperial.ac.uk/media/imperial-college/medicine/sph/ide/gida-fellowships/Imperial-College-COVID19-NPI-modelling-16-03-2020.pdf

6. Robert Verity, Lucy C Okell, Ilaria Dorigatti, Peter Winskill, Charles Whittaker, Natsuko Imai, Gina Cuomo-Dannenburg, Hayley Thompson, Patrick G T Walker, Han Fu, Amy Dighe, Jamie T Griffin, Marc Baguelin, Sangeeta Bhatia, Adhiratha Boonyasiri, Anne Cori, Zulma Cucunubá, Rich FitzJohn, Katy Gaythorpe, Will Green, Arran Hamlet, Wes Hinsley, Daniel Laydon, Gemma Nedjati-Gilani, Steven Riley, Sabine van Elsland, Erik Volz, Haowei Wang, Yuanrong Wang, Xiaoyue Xi, Christl A Donnelly, Azra C Ghani, Neil M Ferguson. Estimates of the severity of coronavirus disease 2019: a model-based analysis. Lancet Infect Dis (2020) Mar 30. pii: S1473-3099(20)30243-7. DOI: https://doi.org/10.1016/S1473-3099(20)30243-7

7. D. Sridhar, M.S. Majumder. Modelling the pandemic. BMJ 369, m1567 (2020). doi: https://doi.org/10.1136/bmj.m1567

8. Giulia Giordano, Franco Blanchini, Raffaele Bruno, Patrizio Colaneri, Alessandro Di Filippo, Angela Di Matteo, Marta Colaneri, Modelling the COVID-19 epidemic and implementation of population-wide interventions in Italy, Nat. Med. (2020) https://doi.org/10.1038/s41591-020-0883-7

9. K. Mizumoto, K. Kagaya, A. Zarebski, G. Chowell. Estimating the asymptomatic proportion of coronavirus disease 2019 (COVID-19) cases on board the Diamond Princess cruise ship, Yokohama, Japan, 2020. Eurosurveillance 25, pii=2000180 (2020); doi: 10.2807/1560-7917.ES.2020.25.10.2000180.

10. S. Zhang, M. Diao, W. Yu, L. Pei, Z. Lin, D. Chen. Estimation of the reproductive number of novel coronavirus (COVID-19) and the probable outbreak size on the Diamond Princess cruise ship: A data-driven analysis. Int J Infect Dis; 93, 201–4 (2020).

11. Censo de Población y Viviendas 2011. Instituto Nacional de Estadistica (Spain). https://www.ine.es. Accessed: March 30, 2020.

12. https://covid19.isciii.es/resources/serie_historica_acumulados.csv. Accessed: April 12, 2020.

13. Giovanna Cowley, Eunice Teixeira Da Silva, Meno Nabicassa, Pedrozinho Duarte Pereira De Barros, Milena Mbote Blif, Robin Bailey, Anna Last. Is trachoma on track for elimination by 2020? Monitoring and surveillance after mass drug administration with azithromycin for active trachoma in Guinea Bissau. BMJ Global Health 2, A62 (2017).

14. Qun Li, Xuhua Guan, Peng Wu, Xiaoye Wang, Lei Zhou, Yeqing Tong, Ruiqi Ren, Kathy S.M. Leung, Eric H.Y. Lau, Jessica Y. Wong, Xuesen Xing, Nijuan Xiang, Yang Wu, Chao Li, Qi Chen, Dan Li, Tian Liu, B.Med Jing Zhao, Man Liu, Wenxiao Tu, Chuding Chen, Lianmei Jin, Rui Yang, Qi Wang, Suhua Zhou, Rui Wang, Hui Liu, Yinbo Luo, Yuan Liu, Ge Shao, B.Med Huan Li, Zhongfa Tao, Yang Yang, Zhiqiang Deng, Boxi Liu, Zhitao Ma, Yanping Zhang, Guoqing Shi, Tommy T.Y. Lam, Joseph T. Wu, George F. Gao, D.Phil Benjamin J. Cowling, Bo Yang, Gabriel M. Leung, Zijian Feng. Early Transmission Dynamics in Wuhan, China, of Novel Coronavirus-Infected Pneumonia. N Engl J Med 382, 1199–1207 (2020). doi:10.1056/NEJMoa2001316.

15. Rebecca Kahn, Corey M. Peak, Juan Fernández-Gracia, Alexandra Hill, Amara Jambai, Louisa Ganda, Marcia C. Castro, and Caroline O. Buckee. Incubation periods impact the spatial predictability of cholera and Ebola outbreaks in Sierra Leone. Proc. Natl. Acad. Sci USA 117, 5067–5073 (2020).

16. Christophe Fraser, Steven Riley, Roy M. Anderson, and Neil M. Ferguson. Factors that make an infectious disease outbreak controllable. Proc. Natl. Acad. Sci. USA 101, 6146–6151 (2004).

17. Corey M. Peak, Lauren M. Childs, Yonatan H. Grad, and Caroline O. Buckee. Comparing nonpharmaceutical interventions for containing emerging epidemics. Proc. Natl. Acad. Sci USA 114, 4023–4028 (2017).

18. J Wallinga and M Lipsitch. How generation intervals shape the relationship between growth rates and reproductive numbers. Proc. R. Soc. B. 274, 599–604 (2007).

19. A. Aleta, Y. Moreno, Y. Evaluation of the potential incidence of COVID-19 and effectiveness of contention measures in Spain: a data-driven approach. *medRxiv*. (2020). doi: 10.1101/2020.03.01.20029801. Available at: https://www.medrxiv.org/content/10.1101/2020.03.01.20029801v2

20. J. Gómez-Gardenes, D. Soriano-Panos, A. Arenas. Critical regimes driven by recurrent mobility patterns of reaction-diffusion processes in networks. Nat Phys 14: 391–5 (2018).

21. A.J. Kucharski, T.W. Russell, C. Diamond, Y. Liu, J. Edmunds, S. Funk, R.M. Eggo, Centre for Mathematical Modelling of Infectious Diseases COVID-19 working group. Early dynamics of transmission and control of COVID-19: a mathematical modelling study. Lancet Infect Dis. 20, 553–558 (2020). doi: 10.1016/S1473-3099(20)30144-4.

22. Qianying Lin, Shi Zhao, Daozhou Gao, Yijun Lou, Shu Yang, Salihu S. Musa, Maggie H. Wang, Yongli Cai, Weiming Wang Lin Yang, Daihai He. A conceptual model for the coronavirus disease 2019 (COVID-19) outbreak in Wuhan, China with individual reaction and governmental action. Int. J. Inf. Dis. 93, 211–216 (2020).

23. Joseph T. Wu, Kathy Leung, Mary Bushman, Nishant Kishore, Rene Niehus, Pablo M. de Salazar, Benjamin J. Cowling, Marc Lipsitch, Gabriel M. Leung. Estimating clinical severity of COVID-19 from the transmission dynamics in Wuhan, China. Nat. Med. 26, 506–510 (2020).

24. Yan Bai, Lingsheng Yao, Tao Wei, Fei Tian, Dong-Yan Jin, Lijuan Chen, Meiyun Wang. Presumed Asymptomatic Carrier Transmission of COVID-19. JAMA 323(14):1406–1407 (2020). doi: 10.1001/jama.2020.2565.

25. Zhen-Dong Tong, An Tang, Ke-Feng Li, Peng Li, Hong-Ling Wang, Jing-Ping Yi, Yong-Li Zhang, Jian-Bo Yan. Potential Presymptomatic Transmission of SARS-CoV-2, Zhejiang Province, China, 2020. Emerg. Infect. Dis 26,1052–1054 (2020).

26. Sean Wei Xiang Ong, Yian Kim Tan, Po Ying Chia, Tau Hong Lee, Oon Tek Ng, Michelle Su Yen Wong, Kalisvar Marimuthu. Air, Surface Environmental, and Personal Protective Equipment Contamination by Severe Acute Respiratory Syndrome Coronavirus 2 (SARS-CoV-2) From a Symptomatic Patient. JAMA 323, 1610–1612 (2020). doi: 10.1001/jama.2020.3227.

27. Luca Ferretti, Chris Wymant, Michelle Kendall, Lele Zhao, Anel Nurtay, Lucie Abeler-Dörner, Michael Parker, David Bonsall, Christophe Fraser. Quantifying SARS-CoV-2 transmission suggests epidemic control with digital contact tracing. Science 10.1126/science.abb6936 (2020) Mar 31. pii: eabb6936. doi: 10.1126/science.abb6936.

28. P.E. Fine. Herd immunity: history, theory, practice. Epidemiol Rev 15, 265–302 (1993).

29. Seth Flaxman, Swapnil Mishra, Axel Gandy, H Juliette T Unwin, Helen Coupland, Thomas A Mellan, Harrison Zhu, Tresnia Berah, Jeffrey W Eaton, Pablo N P Guzman, Nora Schmit, Lucia Callizo, Imperial College COVID-19 Response Team, Charles Whittaker, Peter Winskill, Xiaoyue Xi, Azra Ghani, Christl A. Donnelly, Steven Riley, Lucy C Okell, Michaela A C Vollmer, Neil M. Ferguson, Samir Bhatt. Estimating the number of infections and the impact of non-pharmaceutical interventions on COVID-19 in 11 European countries. Imperial College COVID-19 Response Team Report. (2020). March 30; doi: DOI: 10.25561/77731. Available at: https://www.imperial.ac.uk/media/imperial-college/medicine/mrc-gida/2020-03-30-COVID19-Report-13.pdf

30. J. Riou, C.L. Althaus. Pattern of early human-to-human transmission of Wuhan 2019 novel coronavirus (2019-nCoV), December 2019 to January 2020. Euro Surveillance 25 (4), pii=2000058 (2020) doi:10.2807/1560-7917.ES.2020.25.4.2000058.

